# in-Silico TRials guide optimal stratification of ATrIal FIbrillation patients to Catheter Ablation and pharmacological medicatION: The i-STRATIFICATION study

**DOI:** 10.1101/2024.03.22.24304715

**Authors:** Albert Dasí, Claudia Nagel, Michael T.B. Pope, Rohan S. Wijesurendra, Timothy R. Betts, Rafael Sachetto, Axel Loewe, Alfonso Bueno-Orovio, Blanca Rodriguez

**Author notes:** Corresponding author: Blanca Rodriguez, Albert Dasí, Department of Computer Science, University of Oxford, Wolfson Building, Parks Road OX1 3QD Oxford (UK).

## Abstract

**Background and Aims:** Patients with persistent atrial fibrillation (AF) experience 50% recurrence despite pulmonary vein isolation (PVI), and no consensus is established for second treatments. The aim of our i-STRATIFICATION study is to provide evidence for stratifying patients with AF recurrence after PVI to optimal pharmacological and ablation therapies, through in-silico trials.

**Methods:** A cohort of 800 virtual patients, with variability in atrial anatomy, electrophysiology, and tissue structure (low voltage areas, LVA), was developed and validated against clinical data from ionic currents to ECG. Virtual patients presenting AF post-PVI underwent 13 secondary treatments.

**Results:** Sustained AF developed in 522 virtual patients after PVI. Second ablation procedures involving left atrial ablation alone showed 55% efficacy, only succeeding in small right atria (<60mL). When additional cavo-tricuspid isthmus ablation was considered, Marshall-Plan sufficed (66% efficacy) for small left atria (<90mL). For bigger left atria, a more aggressive ablation approach was required, such as anterior mitral line (75% efficacy) or posterior wall isolation plus mitral isthmus ablation (77% efficacy). Virtual patients with LVA greatly benefited from LVA ablation in the left and right atria (100% efficacy). Conversely, in the absence of LVA, synergistic ablation and pharmacotherapy could terminate AF. In the absence of ablation, the patient’s ionic current substrate modulated the response to antiarrhythmic drugs, being the inward currents critical for optimal stratification to amiodarone or vernakalant.

**Conclusion:** In-silico trials identify optimal strategies for AF treatment based on virtual patient characteristics, evidencing the power of human modelling and simulation as a clinical assisting tool.

## Introduction

Catheter ablation is the most effective therapy for patients with paroxysmal atrial fibrillation (AF)^1^. Since most ectopic triggers originate in the pulmonary veins^2^, pulmonary vein isolation (PVI) is the cornerstone of ablation therapy. Challenges remain for persistent AF, where patients experience 50% recurrence after PVI^3^, suggesting an additional substrate for arrhythmia beyond pulmonary venous ectopic triggers. Nevertheless, since additional empirical ablation has not demonstrated incremental benefit in large clinical trials (STAR AF II^3^ or CAPLA^4^), PVI remains the recommended ablation strategy at the first procedure^5^. In patients who experience AF recurrence despite durable PVI^6^, there is no consensus regarding the optimal further treatment strategy.

The ablation of atrial low voltage areas (LVA, areas of bipolar voltage lower than 0.5 mV in sinus rhythm) has shown superiority to PVI for the first-time in ERASE-AF^7^. However, only 36% of patients recruited had LVA^7^, indicating a limitation to the applicability of LVA ablation. Novel approaches, namely, MiLine^8^ (anterior mitral line) and Marshall-Plan^9^ (sequential elimination of the Marshall bundles, PVI, and right and left atrial lines of block), obtained promising results in preliminary studies. Although apparently effective and approved for testing in larger trials, these strategies rely on extensive ablation lesions, increasing the risk of complications and generation of a pro-arrhythmic substrate. Moreover, empirical application of these strategies is counter to the paradigm of patient-specific ablation of pathological conduction patterns, which has proven very effective in the RECOVER AF study^5^. Thus, the ability to accurately stratify patients with AF recurrence after PVI to those requiring extensive versus minimal substrate modification, those benefitting from empirical versus functional ablation, and those who may be optimally treated only with additional antiarrhythmic drug therapy, would be of tremendous clinical value but remains a major challenge.

Due to their multiscale nature and scalability, human in-silico trials (i.e., clinical trials conducted in large cohorts of virtual patients using human-based computer modelling and simulation) can assist in identifying and explaining individual patient characteristics that guide optimal therapy selection^10,11^. The aim of our i-STRATIFICATION study is to provide evidence for the optimal stratification of patients with AF recurrence after PVI to 13 state-of-the-art pharmacological and ablation therapies, through in-silico trials.

## Methods

The methodology used is explained in detail in the **Supplemental Material** and the files used to conduct simulations are publicly available at https://zenodo.org/records/10562550.

### Overview of study design

A total of 800 virtual AF patients with durable PVI underwent a non-pulmonary vein-based AF induction protocol. The subset with sustained arrhythmia despite PVI were then subjected to 13 independent further treatments for AF (**Figure 1**). This served to identify subgroups of patients that, based on their clinical characteristics, were most likely to respond to a specific treatment. These clinical features were used to inform a decision algorithm for the optimal stratification of AF patients to specific catheter ablation and/or antiarrhythmic drug therapies.

**Figure 1.**
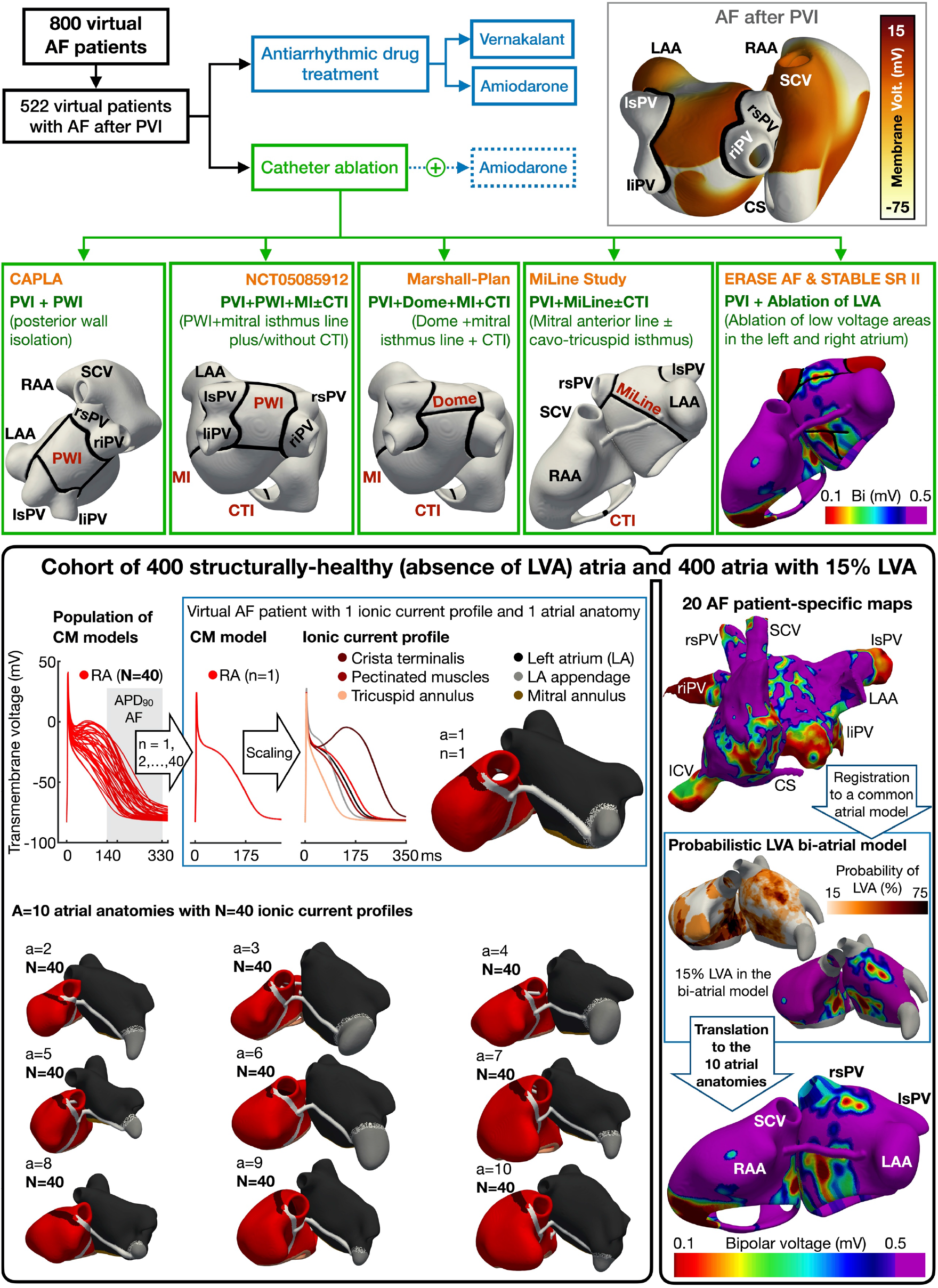
Study design and construction of the cohort of 800 virtual patients. **Top)** Study design: virtual patients with atrial fibrillation (AF) despite pulmonary vein isolation (PVI) are subjected to second procedure treatments. **Bottom)** Population of 400 virtual patients with absence of low voltage areas (LVA): Forty cardiomyocyte (CM) models, representative of right atrial tissue, are considered. The 40 ionic current profiles, with consideration of regional heterogeneities, are used to populate 10 atrial anatomies. Every virtual patient presents a unique combination of ionic current profile and atrial anatomy, resulting in 400 human atria. The cohort is extended to 800 virtual patients by adding LVA into each of the 400 atria, using a probabilistic LVA map derived from 20 AF patients. A representative anatomy with 15% LVA extension in the right and left atrium is illustrated. Abbreviations. **RA-LA**: right and left atrium; **RAA-LAA**: RA and LA appendage; **SCV-ICV**: superior and inferior cava vein; **rs-ri-ls-li-PV**: right superior and inferior, left superior and inferior pulmonary vein; **CS**: coronary sinus.

### Study population

The cohort of 800 virtual AF patients was designed to have variability in anatomy, electrophysiology, and tissue structure, based on human data and recreating the heterogeneity encountered in clinical practice (**Figure 1**). The criteria for the calibration of the population of models using experimental and clinical data are described in full in the Supplementary Material.

In line with STABLE-SR-II^12^, one-half of the cohort was modelled without LVA. These 400 virtual AF patients were developed combining 40 widely-variable ionic current profiles with 10 population-representative atrial anatomies.

#### Anatomical variability

The atrial anatomies in the virtual cohort were selected to span volume ranges associated with AF recurrence (right and left atrial volume ranges of [80, 148] mL and [65, 112] mL, respectively^13^ **Supplemental Figure 1**). Thus, 10 atrial anatomies (with right and left atrial volumes of 127±51 mL and 105±39 mL, respectively; mean ± SD) were derived from a human bi-atrial statistical shape model^14^, based on 47 (40% female) clinical CT and MRI datasets^14^. The anatomies used are described in **Supplemental Table 1**.

#### Electrophysiological variability

Forty atrial cardiomyocyte profiles were identified to capture the variability in action potential characteristics of right atrial trabeculae from 149 (37% female) persistent AF patients^16^. The population of 40 atrial cardiomyocyte models was created by introducing variability in the ionic current densities of a baseline human atrial cellular model^15^ (the scaling factors used are available in **Supplemental Table 2**). Each of these 40 cardiomyocyte models, representative of the right atrial tissue, was scaled to reflect electrophysiological heterogeneities in six atrial regions, as described in **Supplemental Table 3**. Together, the seven action potential models configured a unique ionic current profile (**Figure 1**). Every virtual patient presented a unique combination of ionic current profile and atrial anatomy, resulting in 400 human bi-atrial models with variability in anatomy and electrophysiology. Regional heterogeneities in conduction velocity and anisotropy were also included (**Supplemental Table 3**), setting the baseline plane wave velocity to 80 cm/s in the bulk tissue^11,16^. After considering variability in the ionic current profile (e.g., ±50% variation in I_Na_ density), the population of virtual patients had a longitudinal conduction velocity in healthy tissue (i.e., not defined as LVA) ranging between 58.8 and 102.5 cm/s (86.9±18.3 cm/s; median ± IQR; detailed explanation in the **Supplemental Material**).

#### Structural variability

To develop the other half of the cohort, each of the 400 virtual patients was duplicated into a version that considered LVA. LVA occupied at least 15% of the left atrium, as this LVA extension has been associated with AF recurrence in patient with persistent AF^17^. Moreover, the same LVA degree was modelled in the right atrium, since structural remodelling occurs in both atria^18^.

To incorporate LVA, the bi-atrial electro-anatomical maps of 20 (76% persistent, 41% female) AF patients^19^ were registered to the endocardial surface of the bi-atrial statistical shape model (detailed explanation in the **Supplemental Material**). Then, the bipolar voltage data of all 20 patients were superimposed into the same atrial geometry to build a probabilistic LVA map, i.e., identifying those atrial regions most likely to undergo LVA remodelling. These most probable areas were assigned to be LVA until 15%^17^ of the left and right atrium was remodelled. Finally, the LVA distribution created in the bi-atrial statistical shape model was extended to the 10 atrial anatomies. **Figure 1** shows the 15% bi-atrial LVA extension on a representative anatomy. LVA were simulated as regions of 30% decreased longitudinal conductivity, increased anisotropy (i.e., 8:1 longitudinal to transversal conductivity ratio) and 50%, 40% and 50% reductions in I_CaL_, I_Na_ and I_K1_, respectively^11^. After applying this remodeling on top of the individual electrophysiological properties of each virtual patient, the slowest conduction velocity observed in LVA was 30 cm/s, in agreement with simulation^20^ and clinical studies^21^.

### Pulmonary vein isolation and AF induction

To simulate AF recurrence after PVI, the pulmonary veins of virtual patients were isolated before applying the AF induction protocol. PVI was modelled as in^20^, considering a wide circumferential ablation. For consistency across different atrial anatomies, the circumferential ablation was applied at the minimum distance from the veins that enabled their complete isolation. In order to eliminate the intracellular electrotonic load and closely reproduce ablation lesions clinically, the latter were modelled in-silico by removing the respective elements from the atrial mesh.

After the virtual application of PVI, spiral wave re-entries were imposed in the atria as the initial conditions of the simulation and AF dynamics were analysed for 7 s of activity^11^. This protocol mimicked AF initiation from non-specific triggers outside the pulmonary veins, and a detailed description of the protocol is provided in the **Supplemental Material**.

Simulated 12-lead ECGs were computed in virtual patients with sustained (>7 s) AF. The ECG was computed for 30 s (i.e., duration used clinically for AF diagnosis), to ascertain the proportion of virtual patients that, being initially defined as sustained (>7 s) AF patients, presented uninterrupted arrhythmia over a meaningful duration in the clinics.

### Intervention

Virtual patients with uninterrupted arrhythmia (i.e., > 7 s of organized flutter or AF) after PVI were independently subjected to nine ablation strategies and two antiarrhythmic drugs. Two additional drugs were applied after ablation if arrhythmia sustained for all ablation procedures (**Figure 1**).

The ablation strategies considered: (i) Posterior wall isolation, PWI^4^; (ii) PWI plus mitral isthmus (MI) line, PWI+MI; (iii) PWI+MI plus cavo-tricuspid isthmus line (CTI), PWI+MI+CTI; (iv) Anterior mitral line, MiLine^8^; (v) MiLine+CTI; (vi) Marshall-Plan^9^; and (vii-ix) ablations of LVA in three steps: only isolating LVA in the left atrium, as in STABLE-SR-II^12^ and ERASE-AF^7^, together with CTI block, and with CTI block and right atrial LVA ablation. A drug could be applied on top of the ablation procedure (i.e., synergistic drug and ablation therapy^21^): amiodarone 1.5 μM, as it is recommended for long-term rhythm control of AF^22^.

The pharmacological arm included: (i) amiodarone 3.0 μM; and (ii) vernakalant 30 μM (considered in this study as a potent class IC agent^23^). Drug action was simulated as a simple pore-block model, considering the doses within the therapeutic plasma concentration^11^. **Table 1** shows the percentage of ionic current block at clinically relevant concentrations.

**Table 1.** Ionic current block (%) exerted by the antiarrhythmic drugs.

| Treatment<br>(concentrations) | Ionic current block (%) |  |  |  |  |  |  |  |  | Ref. |
| --- | --- | --- | --- | --- | --- | --- | --- | --- | --- | --- |
|  | I<sub>Kur</sub> | I<sub>Kr</sub> | I<sub>to</sub> | I<sub>K1</sub> | I<sub>Ks</sub> | I<sub>CaL</sub> | I<sub>NaK</sub> | I<sub>NCX</sub> | I<sub>Na</sub> |  |
| Amiodarone (1.5 µM) | - | 40 | - | - | 30 | - | - | 30 | 20 | <sup>11</sup> |
| Amiodarone (3.0 µM) | - | 70 | 30 | 20 | 50 | - | 40 | 50 | 50 | <sup>11</sup> |
| Vernakalant (30 µM) | 70 | 60 | 70 | - | - | 20 | - | - | 70 | <sup>11,23</sup> |

The three-dimensional monodomain equation of the transmembrane voltage and all ECG calculations were solved using the high-performance open-source software MonoAlg3D^24^, with meshes of 400 μm edge length^25^.

## Results

### Primary outcome

Sustained arrhythmia (i.e., organized flutter or AF) occurred in 522 (65%) virtual patients despite PVI (243 and 279 with absence and presence of LVA, respectively; the type of arrhythmia is explained below). All virtual patients with sustained arrhythmia over 7 s also presented uninterrupted arrhythmia for 30 s on the ECG (a representative example is shown in **Figure 2A**). The average ECG dominant frequency was 6.9±3.4 Hz, consistent with clinical measurements^26^.

**Figure 2.**
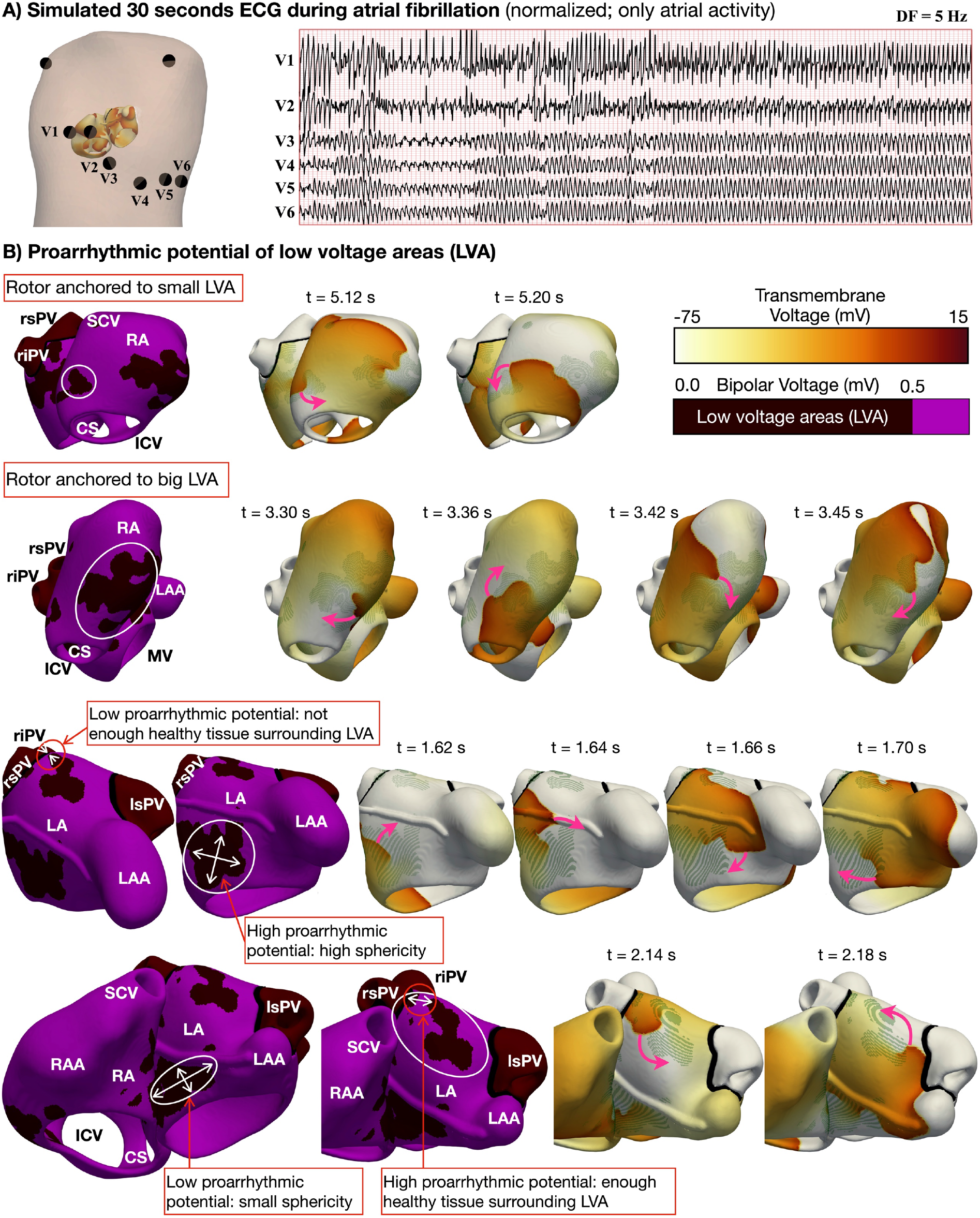
Simulated ECG and proarrhythmic potential of LVA. **A)** Simulated 30 seconds ECG during AF of a representative virtual patient (virtual atrial inside the torso). **B)** Re-entrant drivers anchored to smaller LVA in virtual patients with short refractoriness and slow conduction velocity. Virtual patients with higher conduction velocity showed rotors within bigger LVA. Rotors only anchored to LVA that presented enough healthy myocardium in their surroundings. Higher proarrhythmic potential derived from more spherical rather than elongated LVA shapes. Abbreviations. **RA-LA**: Right and left atrium; **RAA-LAA**: RA and LA appendage; **MV-TV**: Mitral and tricuspid valve; **SCV-ICV**: Superior and inferior cava vein; **rs-ri-ls-li-PV**: Right superior, right inferior, left superior and left inferior pulmonary vein; **CS**: Coronary sinus.

These virtual patients with sustained (>7s) AF despite PVI underwent second treatments. **Table 2** summarizes the efficacy obtained in-silico and in the clinical trial for each treatment.

**Table 2.**
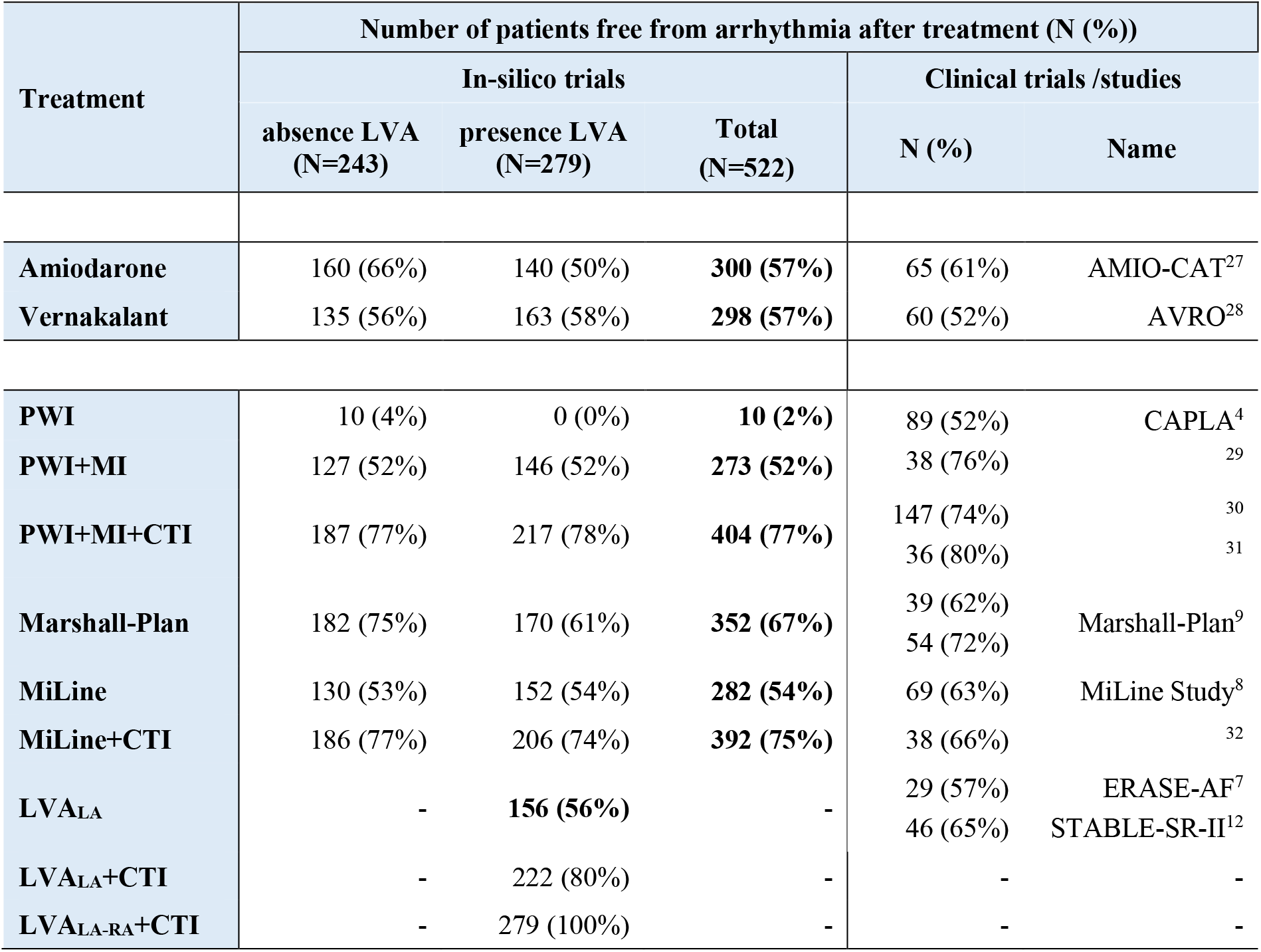
Efficacy of second strategies according to presence of low voltage areas (LVA). Comparison between in-silico and clinical trials. Abbreviations. **PWI**: Posterior wall isolation; **MI**: Mitral isthmus ablation; **CTI**: Cavo-tricuspid isthmus ablation; **MiLine**: Anterior mitral line; **LVA_LA_–LVA_RA_**: Ablation of LVA in the left (LA) and right atrium (RA).

In the cohort of 552 virtual patients with AF after PVI, additional PWI had no incremental benefit, in agreement with CAPLA^4^, and arrhythmia only stopped in 2% of cases. Higher success rates were observed when PWI was applied together with mitral isthmus (PWI+MI) and cavo-tricuspid isthmus ablation (PWI+MI+CTI): 52% and 77%, respectively. Similar efficacies derived from the virtual application of MiLine and MiLine+CTI: 54% and 75%, in agreement with 63-66% reported clinically^8,32^. Virtual Marshall-Plan prevented 67% of AF cases, comparable to 72% observed in human patients^9^. Finally, LVA ablation was applied in the 279 virtual patients with LVA remodelling, and 56%, 80% and 100% efficacy were observed for LVA ablation in the left atrium (LVA_LA_), together with cavo-tricuspid isthmus block (LVA_LA_+CTI) and LVA ablation in the right and left atrium (LVA_LA_+CTI+LVA_RA_), respectively. In the antiarrhythmic drug arm, both amiodarone and vernakalant had a success rate of 57%, as observed clinically^22^. The credibility of in silico trials is supported by the overall agreement with the efficacy reported in clinical trials.

### Arrhythmia despite PVI

In the absence of LVA, 243 virtual patients had inducible arrhythmia despite PVI. In 137 (57%) this was an organised atypical flutter around the rings of the isolated veins, whilst in 106 (43%) it was AF. An average of 2.92±0.62 rotors sustained AF through a wide range of complex dynamics (11% of virtual patients showed only stationary rotors, 70% had one meandering rotor and others stationary, 19% had two or more unstable rotors and 0% presented wave breakups). AF was facilitated by shorter effective refractory period (ERP: 172.3±13.0 vs. 230.6±47.7 ms; virtual patients with AF vs. flutter, respectively; mean ± SD) and larger right atria (i.e., AF proportion increased by 0.21% per mL increase in right atrial volume). The latter enabled sustained arrhythmia in virtual patients with faster conduction velocity (95.0±6.1 vs. 75.3±5.7 cm/s; mean wave velocity in the bulk tissue observed in virtual patients with right atrial volume bigger vs. smaller than 90 mL, respectively).

LVA remodelling increased the proportion of virtual patients sustaining AF to 60% (169 of the 279), whilst the remaining 40% had organized flutter. A higher number of rotors (3.13±0.95) and an increase in AF complexity (18% presented stationary rotors, 43% had one meandering rotor, 32% had two or more unstable rotors and 7% showed wavelets or breakthroughs) owed to a higher amount of pathological conduction patterns located in LVA. **Figure 2B** illustrates representative LVA in the left and right atrium associated with lower and higher proarrhythmic potential.

AF drivers (i.e., sources of electrical activity with the fastest activation time) were observed within LVA only in virtual patients with the shortest ERP (168.5±12.1 ms). When these patients additionally presented slower conduction velocity (75.3±5.7 cm/s), rotors anchored to smaller LVA (**Figure 2B, top**). On the other hand, rotors were observed in bigger LVA (**Figure 2B, middle**) for virtual patients with higher conduction velocity (95.0±6.1 cm/s). Throughout the population, rotors only anchored to dense and more spherical LVA that presented healthy myocardium in their surroundings (**Figure 2B, bottom**). In virtual patients with longer ERP (237.8±46.6 ms), drivers were observed in healthy myocardium, and LVA contributed to AF perpetuation by creating localized irregular activity (i.e., changes of wavefront direction^5^). In these patients, AF maintenance and the success of ablation therapy were determined by the right and left atrial volumes, as discussed below.

### Catheter ablation as second procedure treatment

The success of second ablation procedures was determined by the extent of bi-atrial enlargement and presence of LVA. Thus, positive ablation outcomes derived from targeting anatomic and functional structures, not only in the left but also in the right atrium. Indeed, the importance of additional right atrial ablation is evidenced by comparing the efficacy of strategies that considered CTI ablation (**Table 2**).

#### Additional benefit of cavo-tricuspid isthmus block

Fifty percent of virtual patients presented sustained arrhythmia when only left atrial ablation was considered, regardless of the ablation extent (52%, 54% and 56% arrhythmia freedom after PWI+MI, MiLine and LVA_LA_ ablation, respectively). As explained below, small differences in the efficacy of these strategies regarded whether drivers could still arise in the left atrium post-ablation. Patients with sustained arrhythmia after left atrial ablation were characterized by bigger right atria: no arrhythmia was observed for right atrial volumes lower than 60 mL, and a 40% probability of sustained arrhythmia arose for volumes higher than 100 mL. These patients additionally presented short ERP (172.3±13.0 ms), so that right atrial drivers could still perpetuate AF after extensive left atrial ablation.

On the other hand, 67-80% efficacy derived from additionally considering CTI block (67%, 77%, 75% and 80% arrhythmia freedom after Marshall-Plan, PWI+MI+CTI, MiLine+CTI and LVA_LA_+CTI, respectively). Noteworthy, only 10% of virtual patients where CTI block was successful had originally flutter alone. The other 90% showed a stable macro-re-entry in the right atrium, that potentiated other forms of complex arrhythmias such as unstable rotors anchored to LVA (**Figure 3A**), meandering throughout the right atrial body (**Figure 3B**) or breaking into wavelets (**Figure 3C**). When the macro-re-entry was eliminated through CTI block, unstable rotors spontaneously vanished.

**Figure 3.**
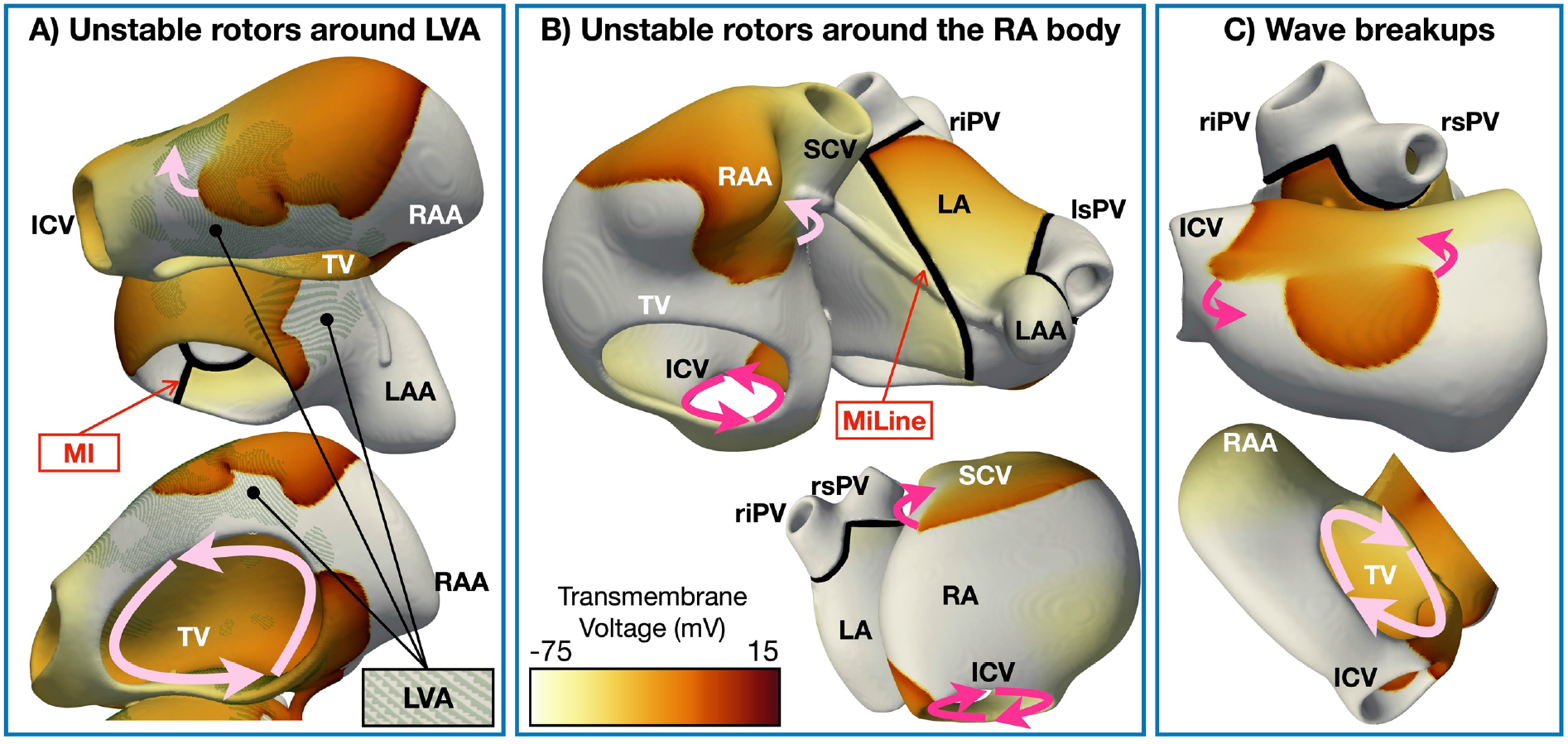
Snapshots of complex arrhythmias sustained by the presence of a macro-re-entrant circuit. The arrows show the direction of rotation. One arrow represents unstable rotors (i.e., rotors that vanish if the macro-re-entry is terminated), and two arrows a macro-re-entrant circuit. Abbreviations. **RA-LA**: Right and left atrium; **RAA-LAA**: RA and LA appendage; **TV**: Tricuspid valve; **SCV-ICV**: Superior and inferior cava vein; **rs-ri-ls-li-PV**: Right superior, right inferior, left superior and left inferior pulmonary vein; **MiLine:** Anterior mitral line of block; **MI:** Mitral isthmus line of block.

When CTI block was applied, the success of left atrial ablation depended on the ablation strategy considered, atrial ERP and presence of LVA. **Figures 4A-4D** illustrate representative examples of unsuccessful ablation for the ablation strategies that considered CTI block. **Figure 4E** shows the number of virtual patients with left vs. right atrial drivers after extensive left atrial ablation. **Figure 4F** separates drivers anchored to LVA from drivers present in healthy myocardium of the right atrium.

**Figure 4.**
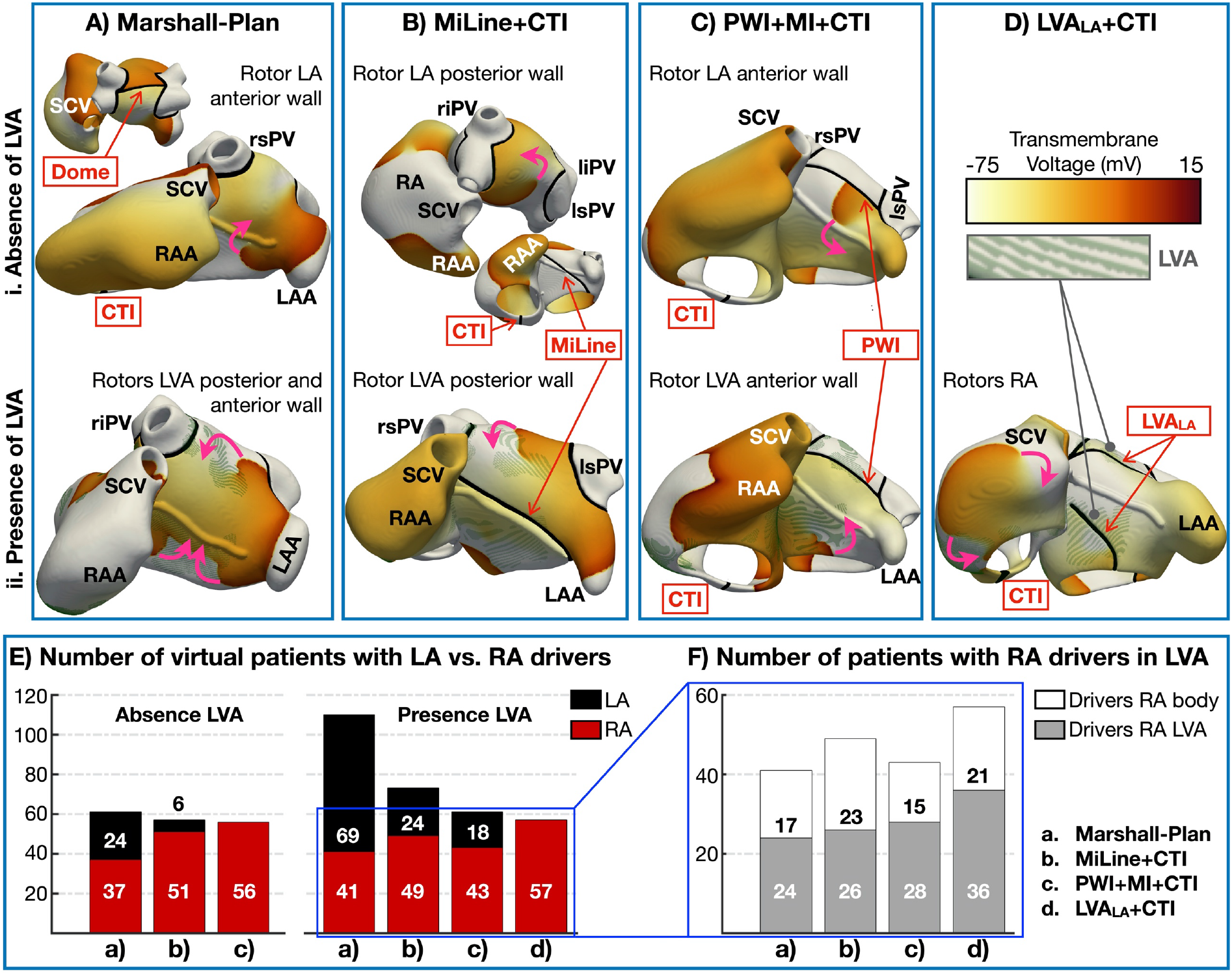
Examples of unsuccessful ablation for the strategies considering cavo-tricuspid isthmus block. **A)** Re-entrant drivers (the arrows show the direction of rotation) in the anterior and posterior wall of the left atrium after Marshall-Plan. **B)** Rotors in the posterior wall after anterior mitral line (MiLine). **C)** Drivers in the anterior wall after posterior wall isolation and mitral isthmus ablation (PWI+MI). **D)** Ablation of low voltage areas (LVA) in the left atrium (LVA_LA_) prevents left atrial drivers, but right atrial drivers may exist. **E)** Number of virtual patients presenting left atrial vs. right atrial drivers after extensive left atrial ablation. **F)** Number of virtual patients with right atrial drivers in healthy myocardium vs. anchored to LVA. Abbreviations. **RA-LA**: right and left atrium; **RAA-LAA**: RA and LA appendage; **SCV-ICV**: superior and inferior cava vein; **rs-ri-ls-li-PV**: right superior, right inferior, left superior and left inferior pulmonary vein.

#### Marshall-Plan

Marshall-Plan had a 75% efficacy in virtual patients with absence of LVA (**Table 2**). The lines of block considered by Marshall-Plan (i.e., dome and mitral isthmus) prevented all arrhythmias in virtual patients with a left atrial volume lower than 90 mL. However, since no ablation lines were applied in the roof or anterior wall, where rotors preferentially located (**Figure 4Ai-ii**), the proportion of AF increased by 0.15% per mL increase in left atrial volume. The efficacy of Marshall-Plan worsened with LVA presence (61%, **Table 2**), since more virtual patients presented drivers anchored to LVA in the left atrial anterior wall (**Figure 4Aii**). In total, 24 and 69 virtual patients with absence and presence of LVA, respectively, presented drivers in the left atrium.

#### MiLine+CTI

Compared to Marshall-Plan, anterior mitral line prevented the formation of rotors in the anterior wall of the left atrium for both patients with absence and presence of LVA. However, after applying MiLine, 6 and 24 virtual patients with absence and presence of LVA, respectively, still presented left atrial drivers. The 6 virtual patients with absence of LVA had a left atrial volume bigger than 140 mL, which facilitated rotor anchoring in the posterior wall (**Figure 4Bi**). The 24 virtual patients with LVA had varying left atrial sizes (100.5±54.5 mL), but all of them presented very short ERP (168.5±12.1 ms). Rotors also appeared in the posterior wall, anchored to small LVA (**Figure 4Bii**). For longer ERP, MiLine was very effective even for large left atria.

#### PWI+MI+CTI

When applied with cavo-tricuspid isthmus block, posterior wall isolation plus mitral isthmus ablation had a similar efficacy in virtual patients with absence and presence of LVA: 77 and 78%, respectively. In the absence of LVA, drivers were mainly located in the right atrium, but unstable rotors appeared in the anterior wall of the left atrium (**Figure 4Ci**). In this subgroup, the proportion of AF increased with increasing right atrial volume (0.16%/mL). Conversely, in the subgroup of virtual patients with LVA, 18 presented stable left atrial drivers in the anterior wall (**Figure 4Cii**).

#### LVA_LA_+CTI

Ablation of LVA prevented all left atrial drivers, showing the highest success rate in the subgroup of virtual patients with LVA. Fifty-seven patients still had sustained arrhythmia, due to right atrial drivers (**Figure 4Dii, 4E**).

#### Arrhythmia despite left atrial ablation and cavo-tricuspid isthmus block

Despite extensive left atrial ablation and CTI block, these four strategies had an efficacy of 67-80% since drivers could still arise primarily in the right atrium (**Figure 4E**). In the subgroup of patients with presence of LVA, drivers were mostly anchored to LVA (**Figure 4F**). Furthermore, even when drivers located in healthy myocardium of the right atrium, LVA helped perpetuating the arrhythmia. Consequently, a prevention of 100% was observed after ablating LVA in the right and left atria plus additional CTI block.

In the subgroup with absence of LVA, 56-61 virtual patients presented right atrial drivers despite left atrial ablation and CTI block (**Figure 4E**). In this subgroup no further ablation was considered, since unstable rotors meandered throughout the right atrial body. Conversely, additional pharmacological treatment successfully stopped the arrhythmias. Low dose amiodarone (i.e., 40% I_Kr_ inhibition) was successful in virtual patients with absence of LVA when applied on top of the ablation.

### Antiarrhythmic drug as second procedure treatment

When antiarrhythmic drugs were virtually-tested post-PVI in the absence of further ablation procedures, the ionic current substrate of the atria determined the response to pharmacological treatment.

#### Amiodarone

Amiodarone prevented all arrhythmia in atria presenting longer ERP (230.7±51.4 ms), resulting from higher I_CaL_ density (0.13±0.08 vs. 0.08±0.02 S/mF; responders vs. non-responders; median ± IQR). For shorter ERP, the success rate of amiodarone was also influenced by the structural substrate (i.e., atrial volume and LVA infiltration). LVA infiltration hampered amiodarone efficacy (66% and 50% for absence and presence of LVA, respectively). Moreover, 88% (73/83) and 82% (114/139) of non-responders with absence and presence of LVA, respectively, were characterized by a bi-atrial volume bigger than 200 mL. Thus, amiodarone was less efficacious in virtual patients with short ERP, big atria and presence of LVA.

#### Vernakalant

Vernakalant increased post-repolarization refractoriness through I_Na_ block and thus, virtual patients with I_Na_ up-regulation were less likely to respond to vernakalant (5.2±1.8 vs. 9.1±1.8 S/mF; responders vs. non-responders). Unlike amiodarone, the efficacy of vernakalant was not influenced by atrial size or LVA infiltration (56% and 58% efficacy for absence and presence of LVA, respectively), but by the excitability (i.e., I_Na_ expression) of virtual AF patients.

### Key patient characteristics and proposed decision algorithm

In an attempt to rationalise the findings of our large-scale in-silico study, **Figure 5** illustrates a possible decision algorithm for optimal stratification to AF therapies based on the assessment of patient characteristics. This algorithm can be evaluated and updated with additional clinical evidence in successive iterations. Based on the simulation results, we propose that the algorithm first evaluates the presence of LVA (i.e., **Figure 5** - right arm of the algorithm), since their ablation yielded the highest prevention efficacy. Virtual patients with LVA and smaller right atria (<60 mL) benefited from LVA ablation in the left atria alone. Conversely, right atrial ablation was required for bigger right atria. Virtual patients with a right atrial volume above 60 mL and short ERP (<170 ms) were optimally treated with LVA ablation in both the left and right atria, with additional CTI block to prevent flutter-derived more complex arrhythmias (Figure 3). In patients with big right atria but longer ERP, CTI block and LVA ablation in the left atrium were sufficient to stop AF maintenance.

**Figure 5.**
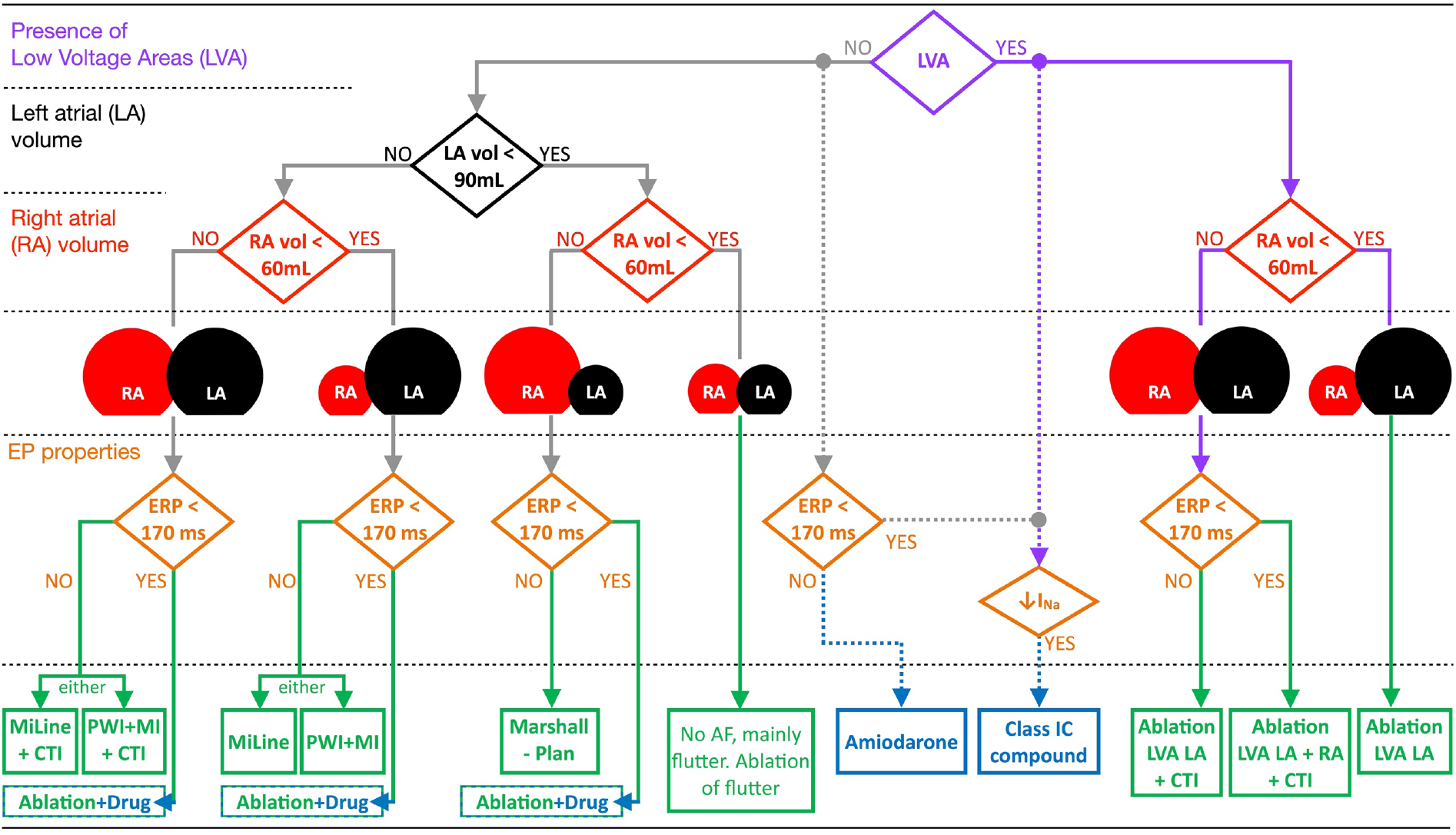
Decision algorithm for optimal stratification of persistent atrial fibrillation patients to catheter ablation or antiarrhythmic drug therapy. Pharmacological treatment is presented with dashed lines since its selection is also influenced by potential concomitant patient’s heart disease^22^. Abbreviations as in Table 2.

In the absence of LVA (i.e., **Figure 5** – left arm of the algorithm), the optimal empirical ablation strategy was also dictated by the atrial volume. Atrial flutter was observed in virtual patients with small right (< 60 mL) and left atria (< 90 mL). Thus, ablation attempted to eliminate the specific re-entrant circuit. For bigger right atria (> 60 mL), additional CTI ablation improved AF prevention. Accordingly, within the strategies considering CTI block, Marshall-Plan was sufficient for small left atria (< 90 mL), but a more extensive ablation, such as MiLine+CTI or PWI+MI+CTI, was required as the left atrial volume increased. In the cases of big left atria and small right atria (< 60 mL), the latter strategies without CTI line (i.e., MiLine and PWI+MI) succeed in preventing AF maintenance.

Noteworthy, virtual patients with absence of LVA but short ERP (< 170 ms) required synergistic pharmacological (i.e., low-dose amiodarone) and ablation therapy. The optimal stratification to ablation strategies was still dependent on the atrial volumes, as with virtual patients with long ERP, but additional antiarrhythmic drugs were needed to prevent AF maintenance.

When administered without catheter ablation, the efficacy of pharmacotherapy was determined by the patient’s ionic profile, being the inward currents (i.e., I_Na_ and I_CaL_) critical for optimal selection of amiodarone or class IC agents (i.e., vernakalant). Due to the current inability to characterise the patient’s ionic current profile on the fly, the stratification to antiarrhythmic drugs in **Figure 5** has been represented with dashed lines (see Discussion).

## Discussion

In this large-scale simulation study, AF maintenance was evaluated in a population of 800 virtual patients presenting variability in anatomy, electrophysiology and tissue structure. In this cohort, 522 virtual patients had inducible arrhythmia after PVI, and were subjected to in-silico trials with ablation and pharmacological treatments (7.064 multi-scale simulations obtained through more than 50,000 hours of computing time). Accordingly, we have demonstrated overall agreement between simulated and clinical results, supporting the credibility of the in-silico trials. Moreover, we have identified specific patient characteristics that dictate treatment success, and provided mechanistic explanations for treatment failure. The digital evidence obtained from the in-silico trials has informed a possible decision algorithm to guide patient stratification to optimal AF treatments. This methodological framework can be probed and updated with further clinical data, to evaluate further treatments for AF, or for other cardiac diseases, highlighting a route for direct translation of human modelling and simulation into clinical practice.

### In-silico trials using modelling and simulation for optimal atrial fibrillation treatment

Computer modelling and simulation has been widely used for testing optimal ablation therapies of AF^33^ in virtual cohorts. Early work by Hwang *et al*.,^34^ showed the feasibility of applying virtual ablation in a cohort of 20 virtual atria reconstructed from CT images of 20 (80% persistent) AF patients. The authors virtually-tested three empirical ablations strategies and, in agreement with our study, concluded that PVI applied together with roof, floor and anterior mitral line had the highest antiarrhythmic efficacy. This work preceded a prospective clinical trial by the same group^35^, in which 108 (78% persistent) AF patients were randomly assigned to receive in-silico guided or standard-of-care ablation. In the in-silico ablation group, the patient’s left atrium was reconstructed and five ablation strategies were assessed. The strategy yielding the fastest AF termination in-silico was selected as optimal therapy and applied in the actual patient. The study obtained non-inferiority of in-silico guided ablation and showcased in-silico trials for guiding the selection of optimal AF therapies.

Further simulation studies^36,37^ included personalization of both the left atrial geometry and the patient-specific fibrotic distribution. Thus, the authors additionally assessed virtual ablation of structurally-remodelled substrate (i.e., LVA or fibrosis derived from late gadolinium-enhancement MRI), which showed higher success rates than empirical ablation. Both studies demonstrated that targeting AF-perpetuating areas (i.e., drivers), yielded the greatest freedom from arrhythmia (i.e., 46%^37^ and 40-80%^36^). In our study, we have also observed the importance of LVA ablation for driver termination. However, after complete isolation of left atrial drivers, we found that right atrial drivers sustained AF in ~40% of cases, not considered in the above-mentioned studies. The importance of right atrial ablation has been illustrated by *Roney et al.*,^38^, in which bi-atrial ablation of fibrosis proved superior to left atrial ablation alone. This was further supported by Boyle *et al*.,^39^ in which the optimal ablation of drivers in both atrial chambers is now being tested in a clinical trial (NCT04101539). In this study, 100 AF patients are expected to undergo simulation-driven ablation, compared to the 800 virtual patients included in our pilot study.

While all these works aimed to develop “digital twins” of the atria for precision medicine, they included no variability in atrial electrophysiology. Failing to consider accurate tissue repolarization properties might lead to believe (among others) that re-entrant AF drivers exclusively localize within regions of structurally-remodelled tissue^36^. In our study, variability in ionic current profiles was considered, resulting in a whole spectrum of AF dynamics (i.e., stable re-entrant drivers anchored to LVA, wave break-ups, unstable and meandering rotors etc.) for the same atrial anatomy and LVA distribution. As shown in this population-based study, including variability in atrial electrophysiological properties is crucial for both accurately phenotyping AF dynamics and properly stratifying patients to optimal ablation and, specifically, pharmacological therapies^10,11^.

The latter concept, patient stratification, is another aspect largely disregarded in both simulation and clinical studies. The “one-size fits all” approach that is currently adopted for rhythm control of AF^40^, and the search for one best treatment, might explain the high recurrence rates reported in persistent AF patients. Conversely, we have shown that the efficacy of AF therapy could be improved if optimal patient stratification, based on key patient characteristics, is considered.

### Efficacy of in-silico vs. clinical trials

Overall agreement between simulated and clinical results was observed, supporting credibility of the in-silico trials. This is important since the results of clinical trials might be confounded by discontinuous monitoring, yielding inflated success, or lack of lesion durability, leading to treatment failure due to gaps in lines rather than ineffective ablation strategy. In this sense, while simulated AF was assessed for 7 seconds (see Limitations), a 100% lesion effectiveness was guaranteed in-silico. Thus, small differences were observed between clinical and simulated results due to procedural differences. For example, most clinical studies allowed concomitant antiarrhythmic drug use or considered first procedure treatments. Therefore, the proposed strategy comprised the efficacy derived from the elimination of pulmonary vein triggers (PVI), plus the added benefit of further lines of block (PVI+). Conversely, our in-silico trials considered second procedure treatments, in which non-pulmonary vein triggers were imposed in every virtual patient. Thus, PVI had no additional benefit, which explained the lower efficacies generally observed in-silico.

This was the case for the virtual application of **posterior wall isolation**. A 52% efficacy was observed in persistent AF patients treated with PWI in CAPLA^4^, as compared to 2% obtained in our study. The posterior wall is believed to be a focus of triggered activity^4^ and thus, the high success of PVI+PWI might be attributed to a complete elimination of ectopy. In our study, we mimicked a situation in which triggers arose everywhere in the atria, which could explain the low efficacy of simulated PWI. Moreover, additional lines of block, such as CTI ablation^29^, are often performed in clinical trials evaluating PWI efficacy, which hampers a direct comparison against in-silico PWI. In agreement with these clinical studies^29,41^, we also observed a high proportion (51%) of virtual patients with atrial flutter after PWI that benefited from additional ablation lines.

Therefore, an important increase in the success rate derived from the virtual application of posterior wall isolation together with **mitral isthmus ablation**. Efficacies of 73.5% and 80% have been reported clinically after PWI+MI in a retrospective^30^ and a small prospective study^31^. In the former study^30^, 30 (15%) persistent AF patients treated with PWI+MI additionally received CTI ablation, and in the latter study^31^, CTI block was applied in 6 (13%) patients. Consistent with the clinical results, we observed 77% efficacy when PWI+MI+CTI was considered.

A similar success rate (75%) derived from the virtual application of **anterior mitral line** and CTI ablation. When MiLine was applied alone, 282 (54%) virtual patients were free of arrhythmia. While a higher efficacy (62.7%) was observed clinically in a cohort of 110 persistent AF patients^8^, 59 of them were treated with additional lines of block (50 with posterior wall isolation and 9 roof line). Moreover, the patients in this study^8^ were carefully selected, so that MiLine was only applied in those presenting large LVA in the left atrial anterior wall. In this case, MiLine regarded a patient-tailored approach^8^, rather than an empirical ablation. Consistent with the clinical study, we observed absence of AF drivers anchored to LVA in the left atrial anterior wall after the virtual application of MiLine.

In this sense, a complete elimination of LVA-dependent drivers derived from **LVA ablation** in both atrial chambers. When LVA ablation was applied only in the left atrium, we observed a 56% success rate, consistent with ERASE-AF^7^. In the latter, from 51 patients with presence of LVA that underwent LVA ablation, 29 (57%) were free from arrhythmia. A slightly higher efficacy (65%) was observed in STABLE-SR-II^12^, where recurrence occurred in 25 out of 71 patients with presence of LVA. This might be attributed to the fact that not all LVA harbour proarrhythmic potential, as shown in this study, and PVI might be enough for preventing arrhythmia.

A slightly higher efficacy was also observed for **Marshall-Plan** in a prospective study^9^ compared to our simulations (72% vs. 67%, respectively). The ablation set of Marshall-Plan was the hardest to reproduce, since the vein of Marshall ethanol infusion could not be accurately modelled in-silico. In VENUS^42^, adding vein of Marshall ethanol infusion proved to reduce recurrence compared to PVI alone, which could explain the slightly lower efficacy of Marshall-Plan observed in-silico. Nevertheless, since we incorporated mitral isthmus ablation, comparable efficacies of 67% and 72% derived from our simulations and clinically^9^. In the prospective study, 79% freedom from arrhythmia was observed in 68 patients presenting a complete lesion set, consistent with the efficacy that we observed in virtual patients with absence of LVA (75%). No subgroup analysis or proportion of LVA is reported in the clinical study for comparison with our simulations.

Likewise for pharmacological therapy, the success rate obtained after virtual administration of amiodarone post-PVI (57%) agrees with the 61% (65/107) reported in AMIO-CAT^27^, a randomized clinical trial assessing whether the short-term use of amiodarone after catheter ablation could prevent early recurrence. The slightly higher efficacy observed in the clinical study^27^ might be due to the fact that PVI only was performed in 75% of patients, with the other 25% receiving additional ablation lines. Overall, the excellent agreement between in-silico and clinical trials is improved when the different procedural aspects are also taken into account.

### Limitations

This study proposes an in-silico trial framework, applied to study 12 AF treatments in a large population of 800 virtual patients. Thus, a compromise was reached between the total number of simulations (i.e., over 7.000) and the time per simulation (i.e., 7 seconds). However, it is unclear whether 7s of simulated AF is representative of the follow-up time reported in clinical trials and thus, if could be used to predict AF recurrence clinically.

Furthermore, we propose a decision algorithm in an attempt to rationalise the large amount of data generated and initiate the process of clinical translation. In its current version, the proposed decision algorithm for patient stratification largely relies on invasive electro-anatomical mapping, even when the algorithm suggests the use of antiarrhythmic drugs without ablation. In this sense, the selection of pharmacological treatment would ideally be guided by non-invasive procedures, even if a redo procedure is common in patients with AF recurrence after PVI. Similarly, the characterisation of the patient’s ionic current profile is not routinely performed in the clinic, due to multiple ethical and procedural reasons. This would impede optimal pharmacological treatment selection according to the in-silico results and thus, other non-invasive biomarkers need to be identified to guide optimal pharmacological therapy. Lastly, while we calibrated and constructed our virtual atria considering at all stages both male and female data, future studies could aim at extending our approach to sex-specific decision algorithms.

## Conclusion

This population-based study highlights the power of in-silico trials based on human modelling and simulation for selecting and understanding optimal cardiac therapies. Simulations demonstrate agreement with clinical results in terms of treatment efficacy, and hold the advantage of identifying specific patient characteristics that dictate treatment success and providing mechanistic explanations for treatment failure. A decision support system is build based on this evidence to guide patient stratification to optimal AF treatments. The validation of the methodological framework and the decision algorithm with further clinical data could boost the efficacy of AF treatment and reinforce the integration of in-silico trials into clinical practice.

## Data Availability

All data produced in the present work are contained in the manuscript

https://zenodo.org/records/10562550

