## Supplemental Material for "in-Silico TRials guide optimal stratification of ATrIal FIbrillation patients to Catheter Ablation and pharmacological medicatION: The i-STRATIFICATION study"

Albert Dasí*, Claudia Nagel, Michael T.B. Pope, Rohan S. Wijesurendra, Timothy R. Betts, Rafael Sachetto, Axel Loewe, Alfonso Bueno-Orovio, Blanca Rodriguez*

* Corresponding author:
Blanca Rodriguez:
Albert Dasí:
Department of Computer Science, University of Oxford, Wolfson Building, Parks Road OX1 3QD Oxford (UK)

### SUPPLEMENTAL METHODS

#### Anatomical Variability

A human bi-atrial statistical shape model[1] was used to generate ten atrial anatomies spanning anatomical variability in clinical data. As illustrated in **Figure S1**, these bi-atrial anatomies covered the volumes associated clinically with atrial fibrillation (AF) recurrence. The ten volumetric bi-atrial geometries are publicly available on (https://zenodo.org/records/5004620), and their corresponding ZENODO identifier is included in **Table S1**.

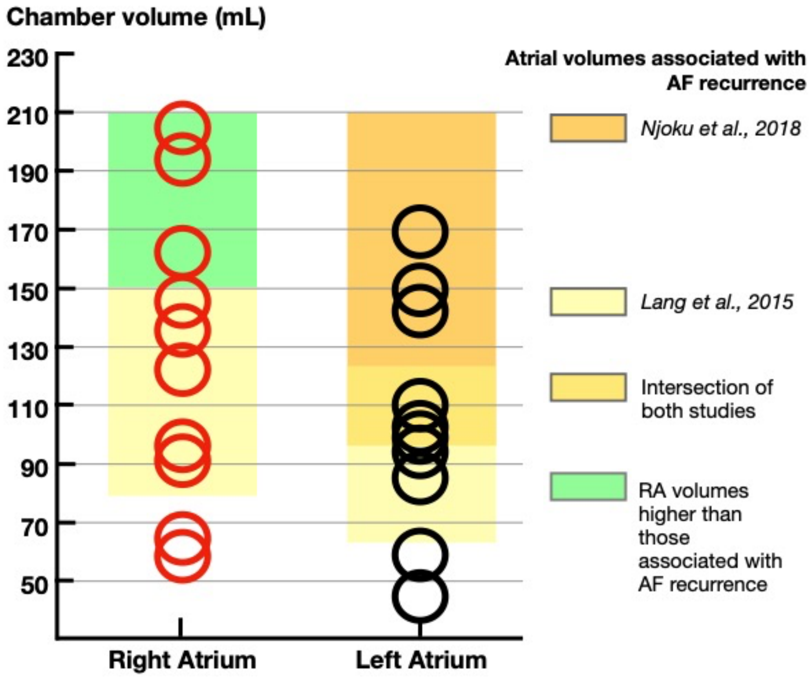


**Figure S1.** Right (RA) and left atrial volumes for the 10 atrial anatomies used in this study for multi-scale simulations. Comparison with the atrial volumes associated with atrial fibrillation (AF) recurrence clinically. The exact volumes for each anatomy are included in **Table S1**.

**Table S1.** Volume of the atrial anatomies considered in this study to conduct multi-scale simulations and corresponding ZENODO identifier. Comparison with the atrial volumes associated with AF recurrence clinically.

| | | ID on ZENODO | Atrial volume (mL) | |
|---|---|---|---|---|
| | | | **Left atrium** | **Right atrium** |
| Atrial anatomy 1 (AA1) | | cn617_g078 | 102 | 58 |
| Atrial anatomy 2 (AA2) | | cn617_g149 | 59 | 64 |
| Atrial anatomy 3 (AA3) | | cn617_g016 | 44 | 91 |
| Atrial anatomy 4 (AA4) | | cn617_g055 | 146 | 96 |
| Atrial anatomy 5 (AA5) | | cn617_g112 | 85 | 122 |
| Atrial anatomy 6 (AA6) | | cn617_g104 | 110 | 135 |
| Atrial anatomy 7 (AA7) | | cn617_g095 | 169 | 145 |
| Atrial anatomy 8 (AA8) | | cn617_g065 | 94 | 162 |
| Atrial anatomy 9 (AA9) | | cn617_g014 | 142 | 194 |
| Atrial anatomy 10 (AA10) | | cn617_g082 | 99 | 205 |
| **Atrial volumes associated with AF recurrence clinically** | Ref [2] | | 65 – 112 | 80 – 148 |
| | Ref [3] | | 96 – 210 | – |

### Mesh transformation for finite volume method (FVM) simulations

Each geometry in ZENODO[1] consists of a VTK file that includes the bi-atrial anatomy with accurate fiber orientation, intra-atrial bridges and material tags in different atrial regions (i.e., right and left atria, right and left atrial appendage, pulmonary veins, superior and inferior cava vein, mitral and tricuspid valve, crista terminalis and pectinated muscles).

The VTK file (i.e., tetrahedral mesh) was converted to an ALG file (i.e., hexahedral mesh), suitable for the GPU solver MonoAlg3D, using the function **hexa-mesh-from-VTK**. This function and a detailed description of its usage can be found in the MonoALG3D GitHub: (https://github.com/rsachetto/MonoAlg3D_C/wiki/Loading-a-VTU-mesh).

For this study, the hexahedral ALG meshes considered a spatial discretization of 400 μm.

### Electrophysiological variability: Population of human atrial cardiomyocyte models

As described in[4], an experimentally-calibrated in-silico population of human atrial cardiomyocyte models was developed using the CRN action potential model[5] as baseline. The methodology has been extensively used in previous studies [4], and consists of scaling a subset of model parameters, which are thought to vary in the human population, over a specified range and retaining the models yielding simulated dynamics as in experiments. By using Latin hypercube sampling, every cardiomyocyte model of the population has a unique combination of parameters, which results in a unique electrophysiological profile and thus, action potential. It is commonly assumed that variability in cellular electrophysiology is mainly expressed at the level of the ion channel density[4].

Building on previous populations studies and given the size of our simulation study, here, an initial population of 60 human atrial cardiomyocyte models was developed by scaling the maximal conductance (G) of key ionic currents over ±50%, including $G_{Kur}$, $G_{Kr}$, $G_{to}$, $G_{K1}$, $G_{CaL}$, $G_{NaK}$, and $G_{Na}$. The generated population, called candidate population of cardiomyocyte models, undergoes a calibration step. The calibration is performed against human experimental data, and ensures that cardiomyocyte models accurately reproduce human electrophysiological properties. For this, action potential recordings from **persistent AF patients**[6] were compared to the in-silico signals. The experimentally-calibrated population comprises those cardiomyocyte models with action potential biomarkers within the ranges delimited by the minimum and maximum experimental value.

From the initial population of 60 cardiomyocytes models, 40 satisfied that all biomarkers (i.e., action potential duration at 90%, 50% and 20% of cellular repolarization, action potential amplitude, resting membrane potential and maximum upstroke velocity) were within the experimental ranges. These 40 cardiomyocyte models, which covered the wide variability in action potential biomarkers from AF patients, were kept for further analysis.

The scaling factors applied to the ionic current densities of the 40 atrial cardiomyocyte models are included in **Table S2**.

**Table S2.** Scaling factors used for developing the in-silico population of human atrial cardiomyocyte models. The population reproduced action potential biomarkers of persistent atrial fibrillation patients.

| Atrial cardiomyocyte model (aCM model) | Scaling factors (no units) applied to the ionic current densities of the CRN model[5] | | | | | | |
|---|---|---|---|---|---|---|---|
| | $G_{Kur}$ | $G_{Kr}$ | $G_{to}$ | $G_{K1}$ | $G_{CaL}$ | $G_{NaK}$ | $G_{Na}$ |
| **aCM model 1** | 0.7621 | 0.6003 | 0.9716 | 0.9698 | 0.5392 | 1.2136 | 1.3596 |
| **aCM model 2** | 0.7504 | 1.2355 | 0.7018 | 0.8079 | 0.7264 | 1.1587 | 0.6016 |
| **aCM model 3** | 0.9618 | 0.6763 | 1.4286 | 1.0725 | 0.5690 | 1.0276 | 0.6278 |
| **aCM model 4** | 0.6867 | 0.8860 | 1.4915 | 0.8829 | 0.5164 | 1.4273 | 0.5102 |
| **aCM model 5** | 0.6629 | 1.4646 | 0.5210 | 1.1076 | 0.8563 | 0.8431 | 1.1721 |
| **aCM model 6** | 0.7321 | 1.0148 | 0.9883 | 1.0247 | 1.0126 | 0.6828 | 1.3770 |
| **aCM model 7** | 0.9306 | 0.7603 | 0.8722 | 1.1521 | 0.9014 | 0.6048 | 0.7826 |
| **aCM model 8** | 1.0910 | 0.9791 | 0.5488 | 1.1223 | 0.6661 | 1.4412 | 0.8503 |
| **aCM model 9** | 1.2735 | 0.9331 | 1.4003 | 1.0963 | 1.1967 | 0.9270 | 1.0428 |
| **aCM model 10** | 1.0626 | 1.3265 | 1.1443 | 0.8156 | 0.5845 | 1.1001 | 1.0654 |
| **aCM model 11** | 1.0131 | 1.4589 | 1.0724 | 0.7910 | 1.3858 | 1.1989 | 0.9874 |
| **aCM model 12** | 0.9409 | 1.3687 | 0.7246 | 0.9926 | 1.0934 | 0.8998 | 1.4146 |
| **aCM model 13** | 1.2338 | 1.4070 | 1.4760 | 0.8378 | 1.0290 | 1.2434 | 1.4282 |
| **aCM model 14** | 0.5568 | 1.1475 | 1.0156 | 0.8991 | 1.3667 | 0.5746 | 0.7015 |
| **aCM model 15** | 1.1624 | 0.6497 | 0.8562 | 0.9085 | 0.6328 | 1.4164 | 0.5587 |
| **aCM model 16** | 1.0299 | 1.2460 | 1.3110 | 1.0664 | 0.8313 | 1.2923 | 1.2811 |
| **aCM model 17** | 0.8396 | 1.3125 | 0.6664 | 1.2132 | 0.7453 | 1.1233 | 0.5301 |
| **aCM model 18** | 1.3928 | 1.1939 | 1.4574 | 0.9511 | 0.9705 | 1.2603 | 1.0369 |
| **aCM model 19** | 1.2830 | 0.8181 | 1.0239 | 1.2453 | 1.1534 | 0.5458 | 0.9741 |
| **aCM model 20** | 1.3513 | 0.7267 | 0.5185 | 1.0047 | 0.6479 | 1.3131 | 0.8766 |
| **aCM model 21** | 0.6079 | 1.2794 | 0.7975 | 1.2292 | 1.2245 | 0.9092 | 0.8853 |
| **aCM model 22** | 1.2124 | 0.8523 | 1.2826 | 1.0386 | 0.9523 | 1.0651 | 0.9224 |
| **aCM model 23** | 0.8656 | 1.0386 | 0.7663 | 1.2397 | 1.2159 | 1.4614 | 1.0830 |
| **aCM model 24** | 0.8137 | 0.5142 | 1.1298 | 1.0473 | 1.0723 | 1.3574 | 0.9466 |
| **aCM model 25** | 0.5210 | 1.1757 | 1.1077 | 0.8224 | 1.3387 | 1.4956 | 0.6640 |
| **aCM model 26** | 1.1030 | 1.4856 | 0.6833 | 0.9106 | 0.8714 | 1.3734 | 0.8180 |
| **aCM model 27** | 1.3045 | 0.8259 | 1.3998 | 1.1198 | 0.7662 | 0.9532 | 1.0095 |
| **aCM model 28** | 0.5940 | 0.9057 | 0.6496 | 0.9894 | 0.7099 | 0.9827 | 0.9188 |
| **aCM model 29** | 0.7025 | 1.2128 | 1.2323 | 1.1833 | 1.4857 | 1.0465 | 0.5837 |
| **aCM model 30** | 0.6440 | 1.3528 | 1.1657 | 0.9376 | 1.4259 | 0.9697 | 1.2034 |
| **aCM model 31** | 1.4437 | 0.7812 | 0.7444 | 1.0811 | 1.1017 | 0.6698 | 0.7655 |
| **aCM model 32** | 0.8400 | 1.0911 | 1.3257 | 1.1454 | 0.8059 | 1.0921 | 0.6533 |
| **aCM model 33** | 1.3791 | 0.8778 | 1.2576 | 1.1339 | 0.9973 | 0.8616 | 1.2558 |
| **aCM model 34** | 0.7953 | 1.0427 | 0.5935 | 0.9469 | 1.3067 | 0.5802 | 0.7532 |
| **aCM model 35** | 1.1314 | 1.1193 | 1.0427 | 1.1787 | 1.4125 | 0.8289 | 0.7308 |
| **aCM model 36** | 0.6323 | 0.9572 | 0.8152 | 1.1604 | 1.4520 | 1.2354 | 1.1120 |
| **aCM model 37** | 1.3375 | 0.7139 | 0.9394 | 0.8701 | 0.6808 | 0.7640 | 0.6850 |
| **aCM model 38** | 0.8817 | 1.3949 | 0.9547 | 0.9778 | 0.6177 | 1.1752 | 1.3395 |
| **aCM model 39** | 1.1482 | 1.2872 | 1.0914 | 1.0529 | 0.8907 | 1.3276 | 1.2363 |
| **aCM model 40** | 1.4638 | 1.4205 | 1.3410 | 0.9297 | 1.2568 | 1.3909 | 1.1526 |

### Electrophysiological heterogeneities in different atrial regions

Each of the 40 cardiomyocyte models, representative of the right atrial tissue, was scaled to reflect electrophysiological heterogeneities in six atrial regions (i.e., left atrium, crista terminalis, pectinate muscles, left atrial appendage and atrio-ventricular rings). The scaling factors are available in **Table S3**. The resulting seven action potential models (i.e., the original cardiomyocyte model and the scaled versions) constituted an ionic current profile. Every virtual patient presented a unique combination of ionic current profile and atrial anatomy.

Regional heterogeneities in conduction velocity and anisotropy were also included, as described in **Table S3**.

**Table S3.** Regional electrophysiological heterogeneities in ionic current densities and conduction velocity, adapted from[7]. The original cardiomyocyte model was included in the right atrial tissue.

| Region | Conductivity ratio Transversal-Longitudinal | CV (cm/s) | $I_{to}$ | $I_{CaL}$ | $I_{Kr}$ |
|---|---|---|---|---|---|
| Right atrium | 1:2 | 80 | 1 | 1 | 1 |
| Left atrium | 1:2 | 80 | 1 | 1 | 1.6 |
| Sinoatrial node | 1:1 | 42 | 1 | 1 | 1 |
| Crista terminalis | 1:10 | 157 | 1.35 | 1.6 | 0.9 |
| Pectinate muscles | 1:2 | 133 | 1.05 | 0.95 | 0.9 |
| Bachmann's bundle | 1:2 | 133 | 1 | 1 | 1 |
| Left atrium appendage | 1:2 | 80 | 0.65 | 1.05 | 2.75 |
| Atrio-ventricular rings | 1:2 | 80 | 1.05 | 0.65 | 3 |

In the baseline CRN action potential model[5] (i.e., with no ionic current density variation) the longitudinal conductivity was adjusted to obtain a baseline plane wave velocity of 80 cm/s in the bulk tissue (i.e., right and left atrium), as shown in **Table S3**. After considering variability in the ionic current profile, especially in $I_{Na}$ density (i.e., main ionic current influencing conduction velocity), the population of virtual patients had a longitudinal conduction velocity in the healthy bulk tissue (i.e., tissue not defined as low voltage areas, LVA) ranging between 58.8 and 102.5 cm/s. Moreover, a similar range of variability was observed in the remaining atrial regions which baseline conduction velocity was different from 80 cm/s (i.e., crista terminalis, pectinate muscles, and Bachmann's bundle). This is in accordance with values of conduction velocity reported clinically[8].

### Structural variability

**Low voltage areas (LVA) registration:** The electro-anatomical maps of 20 (76% persistent, 41% female) AF patients were obtained at the John Radcliffe Hospital in Oxford, by Dr Rohan S. Wijesurendra, Dr Michael T.B. Pope and Dr Timothy R. Betts. All studies were conducted according to the principles of the Declaration of Helsinki, and all patients gave written informed consent.

High density mapping of the atria was performed using an Abbott Advisor HD grid (SE) catheter while pacing at the coronary sinus at a cycle length of 800 ms. The patient electro-anatomical maps were registered to the endocardial surface of the bi-atrial statistical shape model, through a rigid and non-rigid registration.

The registration was performed using MeshMonk[9], which is an open-source toolbox implemented in MATLAB. The program orients, repositions, and scales a template surface (i.e., in this case the patients' endocardial surface) to a target surface (i.e., the endocardial surface of the mean shape model), during a rigid registration step. Subsequently, the template surface is further transformed to fit the specific shape of the target surface, using a non-rigid deformation. To perform the rigid and non-rigid registration, common landmarks need to be placed in the target and template surfaces. For the registration of the patient-specific maps, nine landmarks were placed in the left atrium, namely, the intersection of the four veins with the left atrial posterior wall, the tip of the appendage and four equally-spaced point around the mitral ring. Similarly, 7 points were placed in the right atrial surfaces: the intersection of the superior and inferior cava vein with the venous portion of the chamber, the tip of the appendage and four equally-spaced point around the tricuspid ring.

After registration, the patients' voltage data were subsequently interpolated to the target surface using the nearest neighbor algorithm.

**LVA simulation:** LVAs were simulated as regions of 30% decreased longitudinal conductivity, increased anisotropy (i.e., 8:1 longitudinal to transversal conductivity ratio) and 50%, 40% and 50% reductions in $I_{CaL}$, $I_{Na}$ and $I_{K1}$, respectively[10]. This remodeling was applied on top of the individual electrophysiological properties of each virtual patient. Thus, the 30% reduced longitudinal conductivity and the 40% $I_{Na}$ reduction had a different effect in those virtual patients with the slowest and fastest conduction velocity (i.e., velocity in the bulk tissue of 58.8 and 102.5 cm/s, respectively). After applying these changes in LVA, the slowest conduction velocity observed in LVA was 30 cm/s, in agreement with previous simulation[10] and clinical studies[11].

### AF induction

The AF induction protocol considered arrhythmia as already present in the tissue, and assessed AF maintenance under different patient characteristics. For this, spiral wave re-entries were imposed as the initial conditions of the simulation[12],[13]. Three spiral waves were applied in each atrial chamber. In the left atrium, one spiral wave was induced in the anterior wall, one in the posterior, and another in the inferior wall. In the right atrium, two spiral waves were induced in the venous portion, one around the proximity of the superior cava vein and one close to the inferior cava vein, and a third one in the anterior wall. The direction of rotation was clockwise for two spiral waves in each chamber and counter-clockwise in the third one, ensuring that adjacent re-entries rotated with opposing phase[12].
